# Sustaining Rwanda’s HIV response after elimination of PEPFAR funding: a modeling analysis

**DOI:** 10.1101/2025.04.25.25326450

**Authors:** April D. Kimmel, Zhongzhe Pan, Gad Murenzi, Ellen Brazier, Batya Elul, Benjamin Muhoza, Marcel Yotebieng, Kathryn Anastos, Denis Nash, Central Africa International epidemiology Databases to Evaluate AIDS (CA-IeDEA)

## Abstract

**Introduction:** HIV prevention and treatment supported by the United States President’s Emergency Plan for AIDS Relief (PEPFAR) have saved millions of lives. Rwanda is among the most successful countries around the world in achieving global targets with PEPFAR support. However, abrupt funding uncertainty around PEPFAR raises concerns about continued HIV epidemic control. We projected the impact of the Government of Rwanda’s (GoR’s) capacity to offset the elimination of PEPFAR funding on adult HIV epidemic and care continuum outcomes over 10 years.

**Methods:** Using an HIV policy model calibrated to Rwanda, we assessed: capacity to sustain HIV services at 50% (with no capacity by GoR to cover the PEPFAR funding gap), 75%, 90%, and 100% (with full capacity by GoR to cover the PEPFAR funding gap). Scenarios were operationalized by reducing the number on antiretroviral therapy (ART), with immediate ART discontinuation and proportional decreases in HIV diagnosis, ART initiation, and care re-engagement. We projected HIV epidemic outcomes (HIV prevalence, HIV incidence, number with HIV, new HIV infections, deaths) and care continuum outcomes (percent diagnosed, percent on ART among those diagnosed, percent virally suppressed among those on ART). We calculated differences in projected outcomes for partial or no capacity versus full capacity. Secondary analyses assessed delayed coverage capacity by 1 and 3 years.

**Results:** Compared to full capacity at 10 years, the model projected a 13.9%–38.7% increase in HIV prevalence and 69.0%–246.7% increase in HIV incidence across coverage capacity scenarios. This translated to 29,000–64,000 additional adults with HIV and 20,000–92,000 cumulative new adult HIV infections. Cumulative projected deaths increased by 10,000–51,200. The model projected continual reductions in percent diagnosed at 10 years; percent virally suppressed among those on ART was similar across scenarios. Higher, and more delayed, coverage capacity had projected outcomes similar to lower, and less delayed, coverage capacity.

**Conclusions:** Even in countries like Rwanda that have achieved epidemic control, abrupt and persistent elimination of PEPFAR funding could drastically reverse critical gains. Evidence quantifying the consequences of different capacities to sustain HIV services underscores the high stakes of rapid and sufficient action.

## Introduction

Rwanda is a remarkable global HIV success story, one of only 9 countries worldwide to have achieved the ambitious 95-95-95 UNAIDS targets for epidemic control [1]. Rwanda’s success relies substantially on the commitment of the Government of Rwanda (GoR) through high levels of political will and a multisectoral response and implementation of evidence-based approaches for HIV prevention and treatment [2]. While Rwanda has been moving toward sustainability of its HIV response [3], like many countries in the SSA region, the success of Rwanda’s national response also has hinged on support from external donors, especially the United States President’s Emergency Plan for AIDS Relief (PEPFAR), which funded nearly 40% of the total HIV response and 50% of antiretroviral therapy (ART) in Rwanda in fiscal year 2023–2024 [4, 5] .

The future of PEPFAR, however, remains uncertain. The program was reauthorized by the US Congress for only one year (versus the previous three five-year reauthorizations) until March 25, 2025. And on January 20, 2025, President Trump issued an Executive Order freezing all US foreign aid for 90 days, although PEPFAR subsequently secured a limited waiver allowing some service continuation. An abrupt, persistent elimination of PEPFAR funding will result in an approximately $60-million-dollar funding gap for Rwanda’s national response and setbacks in epidemic control, despite the GoR’s strong willingness to increase domestic funding for HIV programs [6, 7].

Leveraging the cohort study established under the Central Africa International epidemiology Database on AIDS (CA-IeDEA) and the CA-IeDEA Rwanda HIV policy model [8], we projected the impact of the capacity for sustained HIV services, including the GoR’s coverage of the PEPFAR funding gap, on adult HIV epidemic and care continuum outcomes.

## Methods

This study was approved by the Republic of Rwanda National Ethics Committee (approval number RNEC660), Albert Einstein College of Medicine Office of Human Affairs (approval number 2021- 13394) and Virginia Commonwealth University Institutional Review Board (approval number HM20008203). Consent was not obtained, as the study used secondary data with no identifiable information.

### Scenarios

We assessed four scenarios regarding the capacity for sustained HIV services in Rwanda, including the Government of Rwanda (GoR) to cover a 10-year PEPFAR funding gap starting in 2025: *50% sustained* (with no capacity by the GoR to cover the PEPFAR funding gap)*, 75% sustained, 90% sustained,* and *100% sustained* (with full capacity by the GoR to cover the PEPFAR funding gap). All scenarios assumed a complete and persistent elimination of PEPFAR support over 10 years [4, 5]. The scenarios were operationalized in terms of reductions in the total number on ART [9]. Therefore, the *50% sustained* scenario assumed a 50% reduction in the number of people on ART, the *75% sustained* scenario assumed a 25% reduction and the *90% sustained* a 10% reduction. The *100% sustained* scenario assumed no reduction in the number of people on ART and also can be interpreted as no reduction in PEPFAR funding. The scenarios were constant over time.

In secondary analysis, we examined the sensitivity of projected outcomes to the timing of Rwanda’s national response, assuming the GoR’s response may require additional planning such that coverage of the PEPFAR funding gap may not be immediate. Accordingly, we assessed the base case scenarios under the assumptions of 1-year and 3-year temporary delays in response.

### Model

We used the CA-IeDEA Rwanda HIV policy model, a deterministic dynamic compartmental model calibrated to adults 15–64 years [8]. The model includes compartments for HIV disease progression by CD4 stratum and care engagement (i.e., undiagnosed, diagnosed, linked, on ART with suppressed HIV viral load, on ART but not suppressed, lost to follow-up (LTFU)). The modeled population is stratified into 35 sub-populations by age, sex, risk (including female sex workers and men who have sex with men), and urbanicity to capture differences in risk of HIV acquisition. Using 50 best-fitting parameter sets from the calibrated model [8], the model projects HIV epidemic and care continuum outcomes, 2004–2035. For the current analysis, HIV epidemic outcomes include: HIV prevalence (%), HIV incidence (number of new infections per 1,000 population), the annual number of new cases, the number of people with HIV and cumulative deaths among people with HIV. HIV care continuum outcomes include: the percent of people diagnosed with HIV, on ART among those diagnosed, and HIV virally suppressed among those on ART, reflecting global targets. We reported incremental differences and percentage change in mean projected outcomes for the *50% sustained, 75% sustained,* and *25% sustained* scenarios compared, separately, to the *100% sustained* scenario between 2025 and 2035.

### Population distribution and parameter values

We adjusted the model’s 2025 projected population distribution across compartments. For each scenario-specific reduction in ART coverage, we assumed that half of people for whom ART was discontinued in 2025 remained in care with non-ART-related clinical support while the remainder were lost from care. We assumed that people who were virally suppressed, but for whom ART was abruptly discontinued due to elimination of PEPFAR support, had CD4 counts >500 cells/μL [10]; we also assumed that those who were on ART and *not* virally suppressed, but for whom ART was abruptly discontinued due to elimination of PEPFAR support, had CD4 counts >200–350 cells/μL [11].

Model parameter values came from multiple sources (**Table 1**). Parameter values related to HIV acquisition came from the literature and national surveys [12–22]. Transition probabilities for natural history disease progression, HIV viral suppression, virologic failure of available ART regimens, and loss to follow-up came from the CA-IeDEA Rwanda cohort [23]. Transition probabilities for HIV diagnosis and linkage to care came from national surveys and the literature [14–16, 24–27]. We assumed that changes in capacity to sustain HIV programs would decrease probabilities of HIV diagnosis, ART initiation, and re-engagement in care. These decreases reflect potential clinical and resource allocation decisions made due to limited availability of diagnostics, medication, and related infrastructure, as well as potential behavioral changes resulting in delayed initial presentation to clinical care [28, 29]. We modeled the decreases by applying multiplicative adjustments to parameters for HIV diagnosis, ART initiation and re-engagement. These adjustments resulted in a constant number of people on ART for a given scenario over time.

**Table 1.**
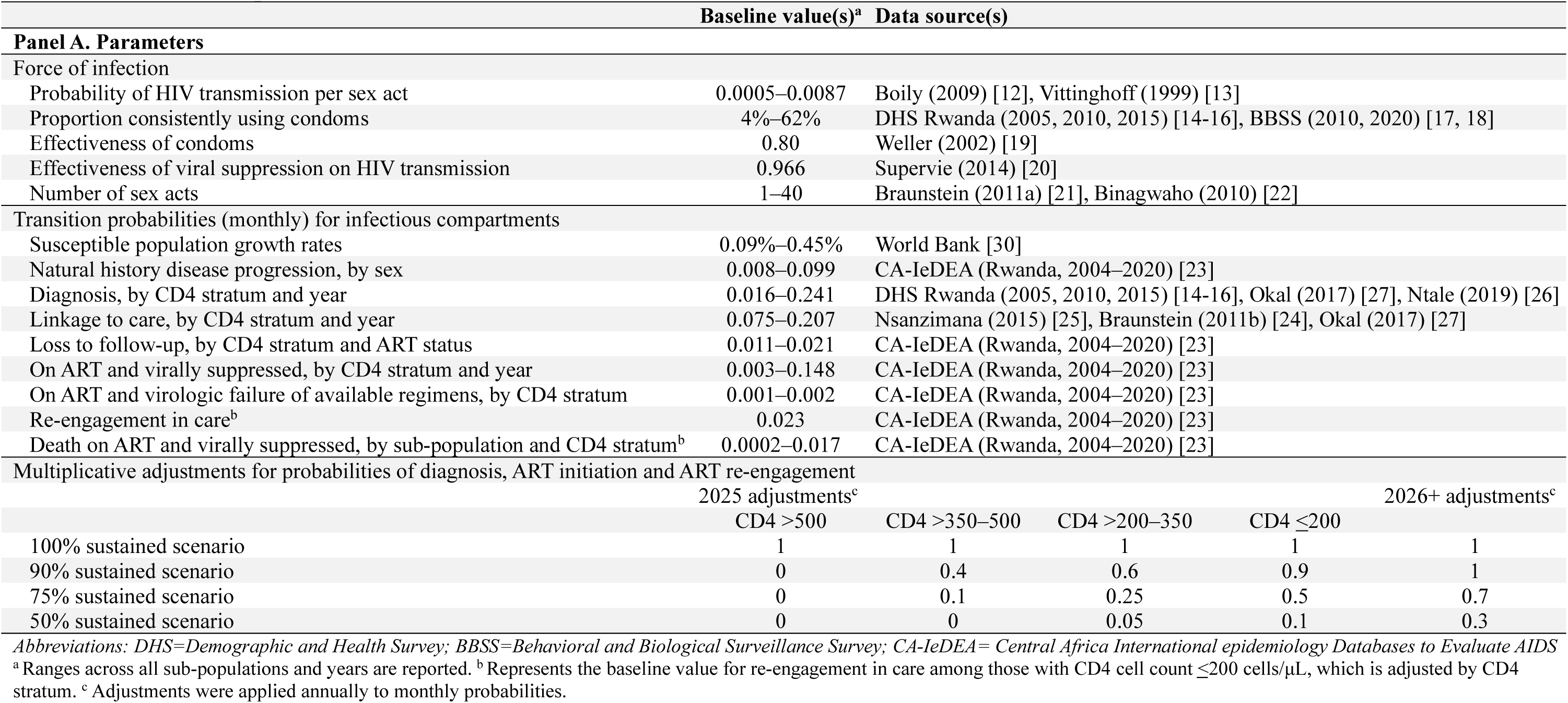
Select model parameters.

## Results

### Baseline analysis

Decreasing capacity for the GoR to cover a complete, persistent PEPFAR funding gap worsened adult HIV epidemic outcomes over time (**Figure 1**). Compared to *100% sustained* at 10 years, projected HIV prevalence increased from 1.8% to 2.0% (+13.9% increase) for *90% sustained* and to 2.5% (+38.7% increase) for *50% sustained.* This was accompanied by projected HIV incidence per 1,000 population increasing from 0.44 to 0.75 (+69.0% increase) for *90% sustained* and to 1.53 (+246.7% increase) for *50% sustained*, with sharp concomitant increases in new HIV cases by nearly 2,800 (+68.3% increase) and 9,900 (+241.9% increase), respectively, at 10 years. Over 10 years, the projected number of adults with HIV increased to over 192,000 (*90% sustained*, +13.8% increase) and to 233,000 (*50% sustained*, +38.0% increase) in 2035. Ten-year cumulative new adult HIV cases increased from 40,000 to 63,000 (*90% sustained*, +58.1% increase) and to 132,000 (*50% sustained*, +229.1% increase), while 10-year cumulative deaths increased to 48,000 (*90% sustained*, +25.9% increase) and to 90,000 (*50% sustained*, +133.8% increase), compared to 38,000 deaths if there were no loss of PEPFAR funding (100% sustained).

**Figure 1.**
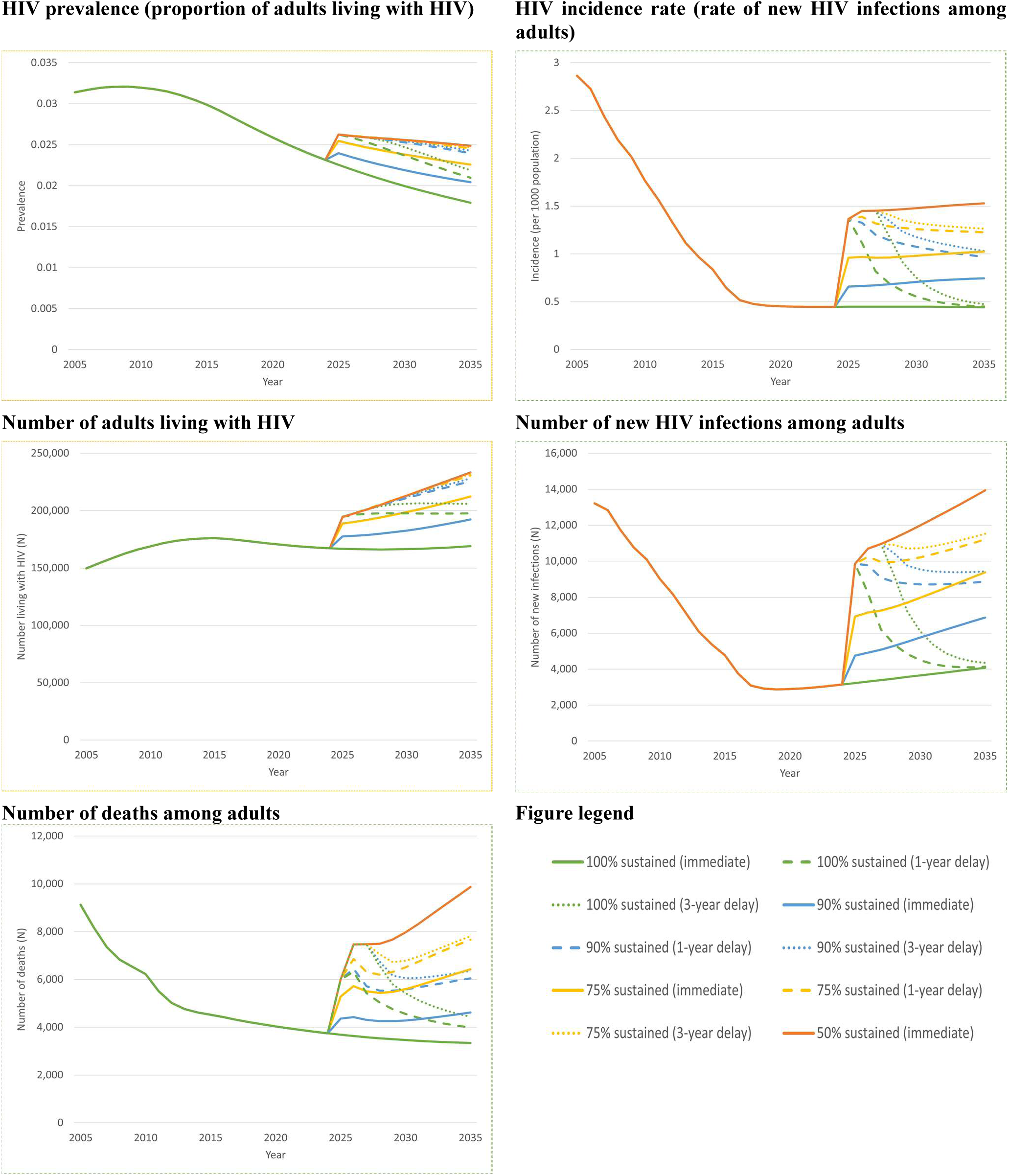
Projected HIV epidemic outcomes in Rwanda by 2035 (ages 15–64 years)

Decreasing capacity by the GoR to cover a complete, persistent PEPFAR funding gap had varied impact on projected HIV care continuum outcomes by 2035 (**Figure 2**). The projected percentage of people diagnosed with HIV had continual reductions, with decreases from 95.3% for *100% sustained* to 79.5% (*90% sustained*, –16.6% decrease) and 53.9% (*50% sustained*, –43.5% decrease) by 2035. There were sharp reductions in the projected percentage of people on ART among those diagnosed in 2025 to as high as a 54.5% decrease (*50% sustained* compared to *100% sustained*). However, these reductions diminished over time: in 2035, projections for ART coverage among diagnosed PLWH decreased from 94.3% for *100% sustained* to 89.6% (*90% sustained*, –5.0% decrease) and to 61.0% (*50% sustained*, –35.3% decrease). Finally, projections for the percentage of people virally suppressed among those on ART by 2035 remained similar across scenarios (approximately a 1% difference), but showed a downward trend.

**Figure 2.**
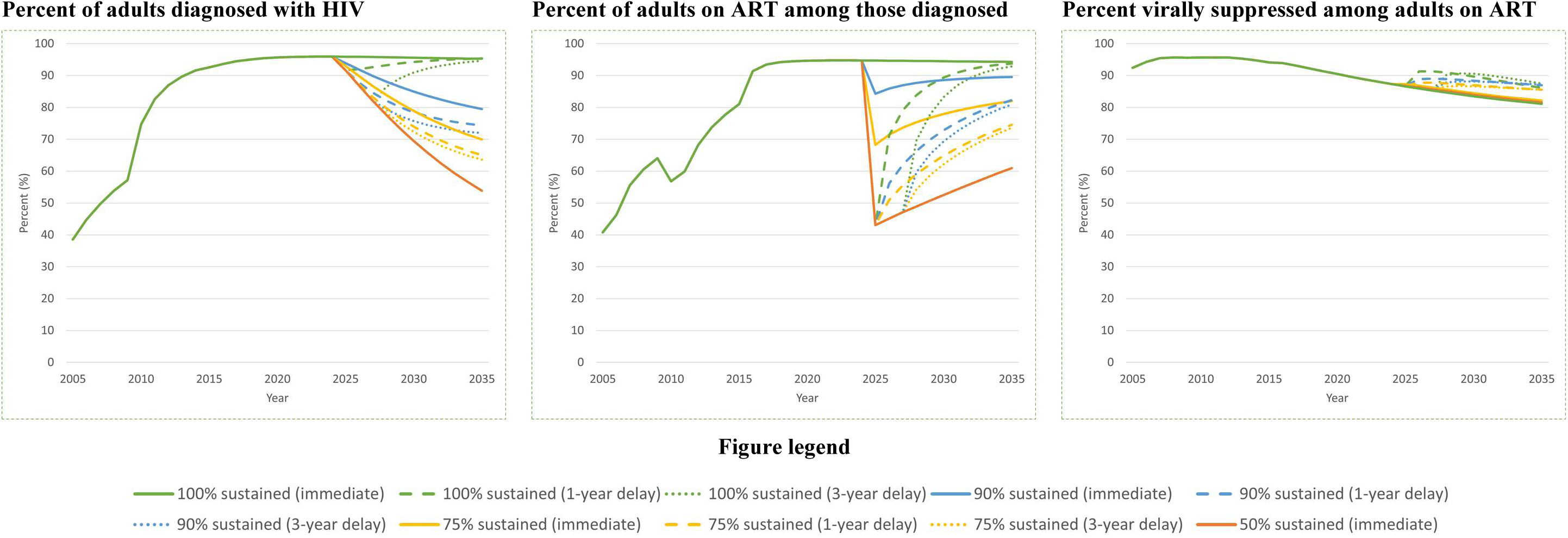
Projected HIV care continuum outcomes in Rwanda by 2035 (ages 15–64 years)

### Secondary analysis

Projected HIV epidemic outcomes were sensitive to the length of time required for GoR to begin covering the PEPFAR funding gap. Across scenarios, projected outcomes generally worsened as delays in covering the PEPFAR funding gap grew, although the impact of the delays diminished as coverage capacity decreased. Further, higher, and more delayed, coverage capacity had projected outcomes similar to lower, and less delayed, coverage capacity. For example, by 2035, the number of new infections projected for a three-year delay in *90% sustained* was similar to immediate *75% sustained* (9,400 vs. 9,400, respectively), while the number with HIV projected from a 3-year delay in 75% sustained was similar to 50% sustained (232,000 vs. 233,000, respectively). Similarly, by 2035, HIV prevalence projected for a one-year delay in *100% sustained* was similar to immediate coverage of *90% sustained*, and the number of cumulative deaths projected for a one-year delay in *90% sustained* was similar to immediate *75% sustained* (26,000 vs.25,000).

For HIV care continuum outcomes, the sensitivity of our findings to delays in coverage capacity varied. Similar to findings for HIV epidemic outcomes, we found that higher, and more delayed, coverage capacity had projected outcomes similar to lower, and less delayed, coverage capacity. For example, by 2035, the percent diagnosed projected for a three-year delay in *90% sustained* was similar to immediate *75% sustained* (72.0% vs 69.9%, respectively), while the projected percent on ART among diagnosed for a one-year delay in *90% sustained* was similar to immediate *75% sustained* (82.3% vs. 82.0%, respectively). Notably, delays in full coverage (*100% sustained*) had minimal impact on percent diagnosed and percent on ART among diagnosed compared to immediate full coverage. However, across all scenarios, the projected percent virally suppressed increased with delays in coverage. Here, the incremental percentage increase in percent virally suppressed was highest for a one-year delay, although this generally decreased as coverage capacity diminished.

## Discussion

Given an abrupt, complete and persistent 10-year withdrawal of PEPFAR funding support, decreasing capacity of the GoR to sustain PEPFAR supported HIV-related programs worsened HIV epidemic outcomes but had varied impact on care continuum outcomes. Delayed coverage of the same coverage capacity worsened all projected outcomes with the exception of full capacity, for which there was minimal impact on some HIV epidemic and care continuum outcomes. Overall, higher and more delayed coverage capacity had projected outcomes similar to lower and less delayed coverage capacity.

Progressively lower capacity to cover the PEPAR funding gap substantially worsened HIV epidemic outcomes over time but had varied impacts on HIV care continuum outcomes. In particular, for HIV care continuum outcomes, there were sharp reductions in the percent of people on ART among those diagnosed in 2025, with these reductions diminishing over time, while the percent of people virally suppressed among those on ART remained similar, although gradually decreasing, across all coverage capacities. The former can be explained by the reductions in the number of people diagnosed with HIV. That is, for people diagnosed with HIV who have either discontinued ART or cannot initiate ART due to resource constraints, the downstream consequences of lack of treatment may include virologic failure, disease progression, and higher risk of developing AIDS-related illness, all of which can lead to increased mortality. This increased mortality, in turn, can reduce the number of people diagnosed with HIV and thus, increase the percent of people on ART among those diagnosed. The latter can be explained by our assumption that the provision of treatment and care for those who were on ART was similar regardless of capacity constraints [31]. The downward trend in the percent virally suppressed among adults on ART across all scenarios reflects the increasing proportion of those on ART who have virologically failed all available ART regimens, highlighting the importance of making available multiple therapeutic drug classes for the treatment of HIV in order to continue to achieve targets even when there are capacity constraints.

Delayed capacity to cover the PEPFAR funding gap worsened projected HIV epidemic and care continuum outcomes, except for the full coverage capacity scenario and one outcome: percent of adults virally suppressed among those on ART. Notably, projected HIV prevalence in the presence of a delay was similar regardless of coverage capacity, except for the full capacity scenario. This could be explained by the immediate and sharp increase in new infections occurring in 2025, which could have long-lasting effects on HIV prevalence and people with HIV. Surprisingly, the projected percent of adults virally suppressed among those on ART increased when there were delays in coverage capacity. This is because, compared to no delays, a greater number of adults for whom ART was discontinued progressed to lower CD4 cell counts. In our model, those with lower CD4 cell counts are more likely to initiate ART than those with higher CD4 cell counts; therefore, a greater number of individuals initiated ART and became virally suppressed, resulting in a higher proportion virally suppressed than without delays in coverage capacity.

Our findings suggest that rapid responses to the PEPFAR funding gap, even if with limited coverage capacity, could have similar or better outcomes than delayed but higher coverage capacity. Indeed, the GoR is increasing its efforts to self-sustain HIV programs and limit the impact of the abrupt HIV financing gap left by PEPFAR. These efforts include increasing government budgets alongside economic development [32], as well as engaging the private sector to sustain the HIV response [4, 33]. With continued effort and political priority of self-sustaining HIV programs, Rwanda has the potential to begin covering the PEPFAR funding gap with limited delay in initiating this coverage. Doing so may provide important insights for other countries in the region who are facing similar situations and mobilizing to respond.

The HIV policy model used in this study was developed and calibrated using Rwanda-specific data from the CA-IeDEA cohort. Long-standing cohort collaborations, such as CA-IeDEA, have played an important role in guiding the HIV response in Rwanda and across the sub-Saharan Africa region. These cohorts provide the real-world, longitudinal data necessary for Ministries of Health to evaluate program impact, identify areas where course corrections may be needed, and plan for sustainability. Leveraging these data to inform policy underscores the importance of cohort-based infrastructure in supporting national decision-making over the coming months and years. Moreover, their use in the current study highlights the importance of supporting Ministries of Health using robust analytic infrastructure—including cohort data platforms—to inform programmatic planning and mitigate the impacts of abrupt transitions to government health programs for large numbers of people.

Our study is among the first to project the impact of sustaining HIV programs on HIV-epidemic and care continuum outcomes, given the elimination of PEPFAR funding. Our results support the concern that abrupt elimination of PEPFAR funding could have significant impact on the progress of ending the HIV epidemic as a public health threat, which had largely been achieved by the GoR, with 50% of its total HIV budget for ART supported by PEPFAR. Our results align with other modeling studies done in the context of low- and middle-income countries (LMICs). In South Africa where PEPFAR funding supported 11% of total HIV budget, a full extraction of PEPFAR funds resulted in 50% increase in HIV incidence in 10 years and reduced percent diagnosed and percent on ART among diagnosed by 3% and 14% in 5 years, respectively [34]. Among 26 LMICs where PEPFAR supports 45% of total HIV spending, a full extraction of PEPFAR funds resulted in 283.3% increase in cumulative new HIV infections and 191% increase in HIV-related death among 26 LMICs over 5 years [35]. These align with our finding that sustaining Rwanda’s HIV budget could result in a 70%–247% increase in HIV incidence, 58%–229% increase in cumulative new infections, 17%–44% reduction in percent diagnosed, and 5%–35% reduction in percent on ART among diagnosed. The differences in magnitude are possibly because our scenarios were defined by restricting only ART availability in Rwanda, which other work assumed either was not affected or affected additional HIV-related programs (e.g., prevention programs) besides ART. In addition, our results of increased cumulative deaths (1,460–5,120 per year) are consistent with a preliminary assessment of excess HIV-related deaths (562 in 1 year) due to pauses in PEPFAR funding [36]. Our projected cumulative deaths are likely higher because we examined a persistent elimination of PEPFAR funding as opposed to a 90-day pause in funding.

Our study has limitations. First, our scenarios focused on decreases in ART coverage only and did not additionally capture impacts of staff shortages to deliver ART and uncertainty in obtaining it. However, we operationalized our scenarios to align with survey findings on the ability to deliver HIV services after extraction of PEPFAR funding, where HIV testing, HIV treatment, and services for LTFU and care re-engagement were among the services most frequently reported cancelled or less active [28]. Second, we did not assess the impact of PEPFAR funding on HIV prevention services (e.g., PrEP) and mother-to-child transmission, because our model structure does not allow for this assessment, and therefore also does not reflect the potential impact on infants, children, and adolescents at risk for and living with HIV. Thus, our projections underestimate the overall impact of PEPFAR funding elimination. Third, we assumed that when the PEPFAR funding gap was covered, HIV testing and ART would be re-initiated at the same rate as prior to the PEPFAR funding elimination. However, extraction of PEPFAR funding may result in health workforce layoffs and closure of HIV-related infrastructure [37], which could have long-lasting impact on HIV service delivery and population outcomes.

## Conclusions

The ability to sustain HIV budgets given the persistent elimination of PEPFAR funding would greatly limit progress to end the public health threat posed by the HIV epidemic and the achievement of 95-95-95 UNAIDS targets in Rwanda. Rapid response to cover the PEPFAR funding gap and to sustain HIV treatment and prevention programs is key to minimizing the effect of PEPFAR funding elimination in Rwanda.

## Competing interests

The authors declare no competing interests.

## Authors’ contributions

A.D.K. and G.M. conceptualized the research. A.D.K. and Z.P. designed the analysis. Z.P. conducted the analysis. A.D.K. and Z.P. drafted the manuscript. A.D.K, Z.P., G.M., B.E., B.M., M.Y., K.A., and D.N. interpretated the data and critically reviewed and revised the manuscript.

## Acknowledgements

Members of Central Africa IeDEA are: Nimbona Pélagie, Association Nationale de Soutien aux Séropositifs et Malade du Sida (ANSS), Burundi; Patrick Gateretse, Jeanine Munezero, Valentin Nitereka, Annabelle Niyongabo, Zacharie Ndizeye , Christella Twizere, Théodore Niyongabo, Centre National de Référence en Matière de VIH/SIDA, Burundi; Hélène Bukuru, Thierry Nahimana, Martin Manirakiza,Centre de Prise en Charge Ambulatoire et Multidisciplinaire des PVVIH/SIDA du Centre Hospitalo-Universitaire de Kamenge (CPAMP-CHUK), Burundi; Patrice Barasukana, Hélène Bukuru, Martin Manirakiza, Zacharie Ndizeye, CHUK/Burundi National University, Burundi; Jérémie Biziragusenyuka, Ella Ange Kazigamwa, Centre de Prise en Charge Ambulatoire et Multidisciplinaire des PVVIH/SIDA de l’Hôpital Prince Régent Charles (CPAMP-HPRC), Burundi; Caroline Akoko, Ernestine Kesah, Esther Neba, Denis Nsame, Vera Veyieeneneng, Bamenda Regional Hospital, Cameroon; Bazil Ageh Ajeh, Rogers Ajeh, Dan Ebai Ashu, Eta Atangba, Christelle Tayomnou Deussom, Peter Vanes Ebasone, Ernestine Kendowo, Clarisse Lengouh, Gabriel Mabou, Sandra Mimou Mbunguet, Judith Nasah, Nicoline Ndiforkwah, Marc Lionel Ngamani, Eric Ngassam, George Njie Ngeke, Clenise Ngwa, Anyangwa Sidonie, Clinical Research Education and Consultancy (CRENC), Cameroon; Anastase Dzudie, CRENC and Douala General Hospital, Cameroon; Djenabou Amadou, Joseph Mendimi Nkodo, Eric Pefura Yone, Jamot Hospital, Cameroon; Annereke Nyenti, Phyllis Fon, Mercy Ndobe, Priscilia Enow, Limbe Regional Hospital, Cameroon; Catherine Akele, Akili Clever, Faustin Kitetele, Patricia Lelo, Kalembelembe Pediatric Hospital, Democratic Republic of Congo; Nana Mbonze, Guy Koba, Martine Tabala, Cherubin Ekembe, Didine Kaba, Kinshasa School of Public Health, Democratic Republic of Congo; Jean Paul Nzungani, Simon Kombela, Dany Lukeba, Sangos plus/Bomoi, Democratic Republic of Congo; Mattieu Musiku, Clement Kabambayi, Job Nsoki,Hopital de Kabinda, Democratic Republic of Congo; Merlin Diafouka, Martin Herbas Ekat, Dominique Mahambou Nsonde, CTA Brazzaville, Republic of Congo; Ursula Koukha, Adolphe Mafoua, Massamba Ndala Christ, CTA Pointe-Noire, Republic of Congo; Jules Igirimbabazi, Nicole Ayinkamiye, Bethsaida Health Center, Rwanda; Providance Uwineza, Emmanuel Ndamijimana, Busanza Health Center, Rwanda; Jean Marie Vianney Barinda, Marie Louise Nyiraneza, Gahanga Health Center, Rwanda; Marie Louise Nyiransabimana, Liliane Tuyisenge, Gikondo Health Center, Rwanda; Catherine Kankindi, Christian Shyaka, Kabuga Health Center, Rwanda; Bonheur Uwakijijwe, Marie Grace Ingabire, Kicukiro Health Center, Rwanda; Beltirde Uwamariya, Jules Ndumuhire, Masaka Health Center, Rwanda; Gerard Bunani, Fred Muyango, Nyagasambu Health Center, Rwanda; Yvette Ndoli, Oliver Uwamahoro, Nyarugunga Health Center, Rwanda; Eugenie Mukashyaka, Rosine Feza, Shyorongi Health Center, Rwanda; Chantal Benekigeri, Jacqueline Musaninyange, WE-ACTx for Hope Clinic, Rwanda; Josephine Gasana, Charles Ingabire, Jocelyne Ingabire, Faustin Kanyabwisha, Gallican Kubwimana, Fabiola Mabano, Jean Paul Mivumbi, Benjamin Muhoza, Athanase Munyaneza, Gad Murenzi, Francoise Musabyimana, Allelluia Giovanni Ndabakuranye, Fabienne Shumbusho, Patrick Tuyisenge, Francine Umwiza, Research for Development (RD Rwanda) and Rwanda Military Hospital, Rwanda; Jules Kabahizi, Janviere Mutamuliza, Boniface Nsengiyumva, Ephrem Rurangwa, Rwanda Military Hospital, Rwanda; Eric Remera, Gallican Nshogoza Rwibasira, Rwanda Biomedical Center, Rwanda. Adebola Adedimeji, Kathryn Anastos, Jean Claude Dusingize, Lynn Murchison, Viraj Patel, Jonathan Ross, Marcel Yotebieng, Natalie Zotova, Albert Einstein College of Medicine, USA; Ryan Barthel, Ellen Brazier, Heidi Jones, Elizabeth Kelvin, Denis Nash, Saba Qasmieh, Chloe Teasdale, Institute for Implementation Science in Population Health, Graduate School of Public Health and Health Policy, City University of New York (CUNY), USA; Batya Elul, Columbia University, USA; Xiatao Cai, Don Hoover, Hae-Young Kim, Chunshan Li, Qiuhu Shi, Data Solutions, USA; Kathryn Lancaster, The Ohio State University, USA; Mark Kuniholm, University at Albany, State University of New York, USA; Andrew Edmonds, Angela Parcesepe, Jess Edwards, University of North Carolina at Chapel Hill, USA; Olivia Keiser, University of Geneva; Stephany Duda; Vanderbilt University School of Medicine, USA; April D. Kimmel, Virginia Commonwealth University School of Public Health, USA.

## Funding

Research reported in this publication was supported by the U.S. National Institutes of Health’s National Institute of Allergy and Infectious Diseases, the *Eunice Kennedy Shriver* National Institute of Child Health and Human Development, the National Cancer Institute, the National Institute of Mental Health, the National Institute on Drug Abuse, the National Heart, Lung, and Blood Institute, the National Institute on Alcohol Abuse and Alcoholism, the National Institute of Diabetes and Digestive and Kidney Diseases, and the Fogarty International Center under Award Number U01AI096299 (Central Africa IeDEA). Informatics resources are supported by the Harmonist project under Award Number, R24AI24872.

## Data Availability Statement

The data used to populate the policy model and that support the findings of this study are available in Table 1 and in the supplementary content of Kimmel et al. Development and calibration of a mathematical model of HIV outcomes among Rwandan adults: informing achievement of global targets across sub-populations in Rwanda, Forthcoming from PLOS One.

## Notes

### Competing Interest Statement

The authors have declared no competing interest.

### Author Declarations

This study was approved by the Republic of Rwanda National Ethics Committee (approval number RNEC660), Albert Einstein College of Medicine Office of Human Affairs (approval number 2021-13394) and Virginia Commonwealth University Institutional Review Board (approval number HM20008203).

